# Pilot proteomics study suggests elevated tumorigenesis-promoting biomarkers in healthy adults after air pollution exposure

**DOI:** 10.1101/2025.09.12.25335654

**Authors:** Victoria Y. Ding, Di Lu, Yinyao Ji, Mary M. Johnson, Francois Haddad, Juyong B. Kim, Benjamin D. Horne, Jin-Ah Park, Kari C. Nadeau, Manisha Desai, Holden T. Maecker

**Author notes:** **Address for Correspondence:** Holden T. Maecker, PhD, Stanford School of Medicine, Fairchild Science Bldg, D039, 299 Campus Drive, Stanford, CA 94305.

## Abstract

**Objectives:** Recent studies have shown that immune and cardiometabolic biomarker levels increased in individuals exposed to ambient air pollution. We sought to explore the feasibility of identifying a pollution exposure signature using the Olink Explore platform in a small, pilot cohort of firefighters and age-matched controls in the Bay Area.

**Methods:** Demographics and plasma samples were collected at two study visits, one following the resolution of California’s record-setting 2020 wildfires and another in the spring of 2021. Age-matched samples were processed on the same plate, with 10 repeated on a second plate for assessing inter-plate variation. For comparing firefighter profile with healthy controls, we analyzed Olink proteomics data under multiple approaches for feature selection, including LASSO (the least absolute shrinkage and selection operator), elastic net, and PLS-DA (partial least squares discriminant analysis). In addition, we selected 67 biomarkers related to well-defined pathways for targeted analyses, whereas 1463 unique assays were considered for untargeted analyses.

**Results:** Our analysis found that levels of angiopoietin 1 and hydroxyacylglutathione hydrolase were consistently higher in firefighters compared to controls. Moreover, matrix metalloproteinase-1, a protein that promotes tumorigenesis, was higher at the visit with greater acute exposure. Fifteen other key biomarkers were jointly identified under untargeted and targeted approaches. When comparing targeted biomarkers in firefighters by level of exposure using a mixed effects regression model, 10 were found to be numerically higher in firefighters with high exposure.

**Conclusions:** These analyses demonstrate the potential utility of large-scale plasma proteomics in identifying a pollution exposure signature.

## Introduction

Wildfires have intensified in the past decade, resulting in lasting social and economic impact. Additionally, fine particulate matter and other toxicants from wildfire emissions have been associated with a range of respiratory and cardiovascular issues,^1^ while their immunologic effects remain underexplored. In our previous study, plasma samples collected during or immediately after the 2014 El Portal wildfire revealed that forest fire exposure in a 12-to 25-year-old cohort was associated with higher levels of interleukin-1β and C-reactive protein compared to age-matched non-smoke-exposed controls, suggesting a proinflammatory state after acute exposure.^2^ The more recent 2020 wildfire season—the worst in California’s history and burning over 4.3 million acres^3^—furthered the opportunity to pilot novel technology and methods for investigating the impact of wildfire pollution on proteomic transducers of health.

Dimensionality reduction techniques, such as partial least squares discriminant analysis (PLS-DA), have traditionally been used in omics studies, although sparsity-promoting methods for biomarker discovery—notably the least absolute shrinkage and selection operator (LASSO)— have gained momentum. Using both classical and novel approaches, the present study seeks to identify a pollution exposure signature in firefighters, as well as to assess differences over time and by level of exposure. Biomarkers targeting well-identified pathways were gleaned from current literature, and we hypothesized that levels of inflammasome markers would be higher in firefighters compared to age-matched unexposed controls. This research adds to the expanding evidence on the health effects of wildfire smoke exposure and enhances the methodological resources available for investigating related issues.

## Methods

### Study population and data processing

Our study included active-duty firefighters and healthy civilian adults residing in the Bay Area, California. Unexposed civilians were age-matched to firefighters in case-control analyses, and exposed participants were considered in longitudinal analyses. Firefighters provided blood samples at two time points, from March to June 2021. Unmatched controls were also assessed twice over the course of one year, from October 2019 to August 2020. Matched controls were drawn from 2017-2021 cohorts and assessed once. Demographics and clinical characteristics were collected on all participants. Level of exposure, as determined by the number of structural fires, was recorded for firefighters only. Samples were processed and stored according to published methods.^4^

Plasma samples were processed using Olink Explore, a high-multiplex, high-throughput protein biomarker platform that measures the relative concentration of proteins in liquid biopsies using next generation sequencing and operating under a semi-automated protocol. Up to 1536 assays are generated across four panels—Cardiometabolic, Inflammation, Neurology, and Oncology. The assay was performed at the Stanford Genomics Center, according to protocols supplied by the vendor (Olink, ThermoFisher, Upsalla, Sweden). Per Olink Explore’s built-in process, each sample measurement was normalized to an internal control and log2 transformed. The median of the plate controls was then subtracted, and resulting values were intensity-normalized (i.e., autoscaled) to achieve mean 0 and standard deviation (SD) 1 for all markers. The intensity-normalized data were used for feature selection, whereas the log2 transformed data were used for assessing inter-plate variability.

Samples from firefighters and their matched controls were run on one plate, and samples from unmatched controls were run on a second plate. Ten samples were run on both plates to assess inter-plate variability.

Targeted biomarkers were selected from the Olink panel based on previous studies investigating biomarkers of pollution exposure and biomarkers associated with survival and major cardiovascular outcomes from large epidemiology studies.

### Statistical analysis

Participant characteristics were summarized by case-control status. In primary analysis, pollution exposure signatures in firefighters were identified under both targeted and untargeted approaches in comparison to the matched control group. Under each approach, we used *t*-tests with false discovery rate (FDR) controlled at 15% to assess marker-by-marker differences between firefighters and matched controls, as well as three multivariable techniques—the LASSO,^5^ elastic net,^6^ and PLS-DA.^7^ Notably, the LASSO has been shown to yield sparse solutions compared to classical dimensionality reduction techniques,^8^ thereby allowing us to identify features with the largest and most consistent signal for future study. The LASSO is particularly effective in high-dimensional settings and helps to prevent overfitting by imposing a penalty on the regression coefficients. In our analyses, the value of the penalty parameter, λ, that minimized the binomial deviance was determined using 10-fold cross-validation. For elastic net, joint cross-validation for λ and the mixing parameter, α, was implemented using the ‘caret’^9^ workflow, which invokes the ‘glmnet’^10^ R package.

PLS-DA is an extension of principal components analysis that additionally considers the correlation between the dependent and independent variables when transforming the original data, i.e., “projection to latent structures.” Its sparse variant, sPLS-DA, used in our analysis prioritizes selection of the most discriminative features for sample classification.^11^ We used 3-fold cross-validation repeated 50 times to determine the optimal number of components to retain and applied the variable importance in projection (VIP) > 1 criterion. The number of variables to select on each component and the prediction distance to evaluate the classification and prediction performance of sPLS-DA were also tuned. These steps were implemented using the ‘mixOmics’^12^ R package. For assessing predictive performance of multivariable methods, AUC with 95% confidence interval were estimated using the ‘pROC’^13^ R package.

In secondary analyses, we separately evaluated targeted marker stability in firefighters and unmatched controls over two time points using paired *t*-tests, with unadjusted p-values meant to be descriptive. Among firefighters, we also explored differences in biomarkers by exposure level (high vs. low, dichotomized based on number of fires). Comparisons between high and low exposure groups were performed in the mixed effects model framework to utilize both time points and account for confounding by age. For each marker, the model included fixed effects of exposure status (high vs. low) and visit (1 or 2), and within-subject correlation was incorporated via a random effect. Estimated exposure group differences with 95% confidence intervals were presented as a forest plot for all targeted markers.

For quality control, we considered all markers included in Olink Explore 1536, as well as our curated list of 67 target markers. Consistent with Olink’s assay validation approach, we estimated Spearman’s rank correlation coefficient (a nonparametric measure of monotonic association, with values closer to ±1 indicating greater strength of rank correlation) for the subset of samples processed on multiple plates, with strength of correlation interpreted according to conventional thresholds.^14^ All analyses were performed in the R statistical computing environment, version 4.3.^15^

## Results

Baseline characteristics of matched participants (median age 41; 56% Non-Hispanic White) are summarized in **Table 1**, with standardized mean differences small (i.e., <0.5) across sex, ethnicity, and allergy history. Unmatched controls were younger (median age 30) and otherwise similar to matched controls.

**Table 1.**
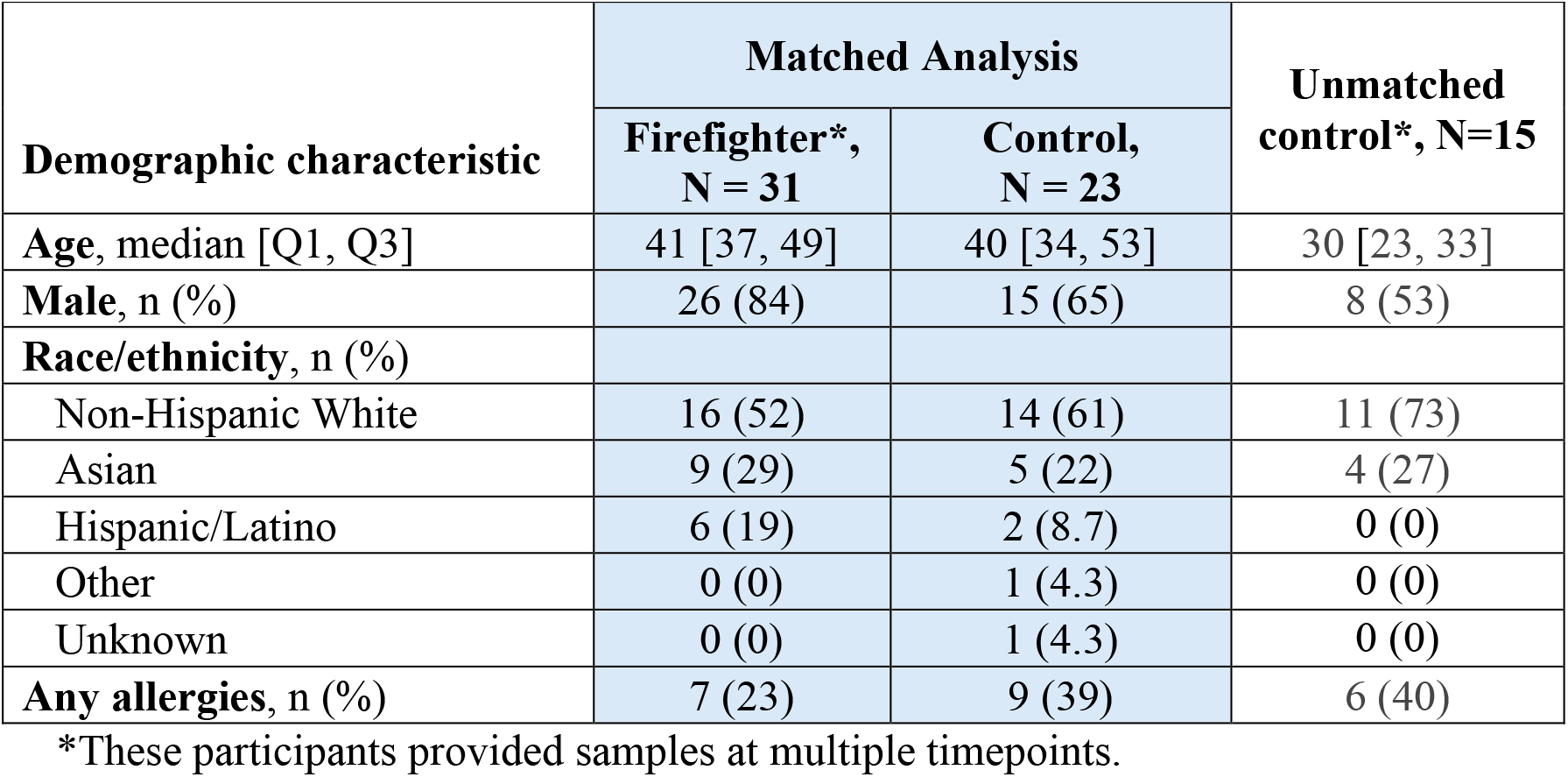
Characteristics of participants with samples processed using Olink Explore.

When considering samples collected at the initial visit, 22 biomarkers were selected by the LASSO, compared to 95 by PLS-DA and 294 by elastic net. Follow-up samples yielded 26 biomarkers selected by the LASSO, compared to 93 by PLS-DA and 158 by elastic net. The LASSO remained as expected as or more parsimonious than the other methods in targeted selection (4 vs. 10 and 14 at the initial visit and 9 vs. 9 and 29 at the follow-up visit). The 18 biomarkers concurrently selected by all multivariable methods, by visit and approach, are summarized in **Table 2**.

**Table 2.** Markers selected by all multivariable methods, by study visit.

| Olink Explore markers, by panel | Initial visit |  | Follow-up visit |  |
| --- | --- | --- | --- | --- |
|  | Targeted | Untargeted | Targeted | Untargeted |
| **Cardiometabolic** |  |  |  |  |
| G protein-coupled receptor 37 |  | * |  | * |
| Granzyme H‡ | * | * | * |  |
| Pleiotrophin |  |  |  | * |
| Surfactant protein D‡ | * | * |  |  |
| **Inflammation** |  |  |  |  |
| Angiopoietin 1 |  |  |  |  |
| Interleukin-16‡ |  |  | * |  |
| Matrix metalloproteinase-1 |  |  |  |  |
| Nibrin‡ |  |  | * |  |
| Neutrophil cytosolic factor 2 |  |  |  |  |
| PC4 and SFRS1 interacting protein 1 |  | * |  |  |
| **Neurology** |  |  |  |  |
| CD8 alpha |  | * |  |  |
| Cytotoxic and regulatory T cell molecule |  | * |  |  |
| Peroxiredoxin 1 |  |  |  | * |
| T-cell leukemia/lymphoma 1A |  |  |  | * |
| **Oncology** |  |  |  |  |
| Beta-defensin 4A and 4B |  |  |  | * |
| Fibroblast growth factor 21 |  | * |  |  |
| Glucagon |  | * |  |  |
| Hydroxyacylglutathione hydrolase |  |  |  |  |

‡Marker on targeted list, as visualized in Figure 1.
Note: Cells of selected markers are shaded in red if relative abundance was higher in firefighters compared to matched controls and in blue if relative abundance was lower. Significant differences under univariate comparisons with the false discovery rate controlled at 15% using the Benjamini-Hochberg procedure are denoted by an asterisk.

At both visits, angiopoietin 1 (ANGPT1) and hydroxyacylglutathione hydrolase (HAGH) were consistently higher in firefighters compared to controls. At the follow-up visit with greater exposure to structural fires on average, matrix metalloproteinase-1 (MMP1) was also higher in firefighters. All but one of the remaining biomarkers were significantly lower in firefighters in univariate, FDR-corrected comparisons.

In the targeted biomarker analysis under a mixed effects regression framework, nine biomarkers showed significantly lower levels in the high exposure group, including macrophage migration inhibitory factor (MIF), granzyme H, and myeloperoxidase (MPO). Additionally, 10 biomarkers exhibited elevated levels in highly exposed firefighters, though these differences did not reach statistical significance. Among these, insulin-like growth factor binding protein 1 (IGFBP1), N-terminal pro-brain natriuretic peptide (NT-proBNP), and surfactant protein D (SFTPD) showed the largest age-adjusted differences. All age-adjusted exposure differences are summarized in **Figure 2**.

**Figure 1.**
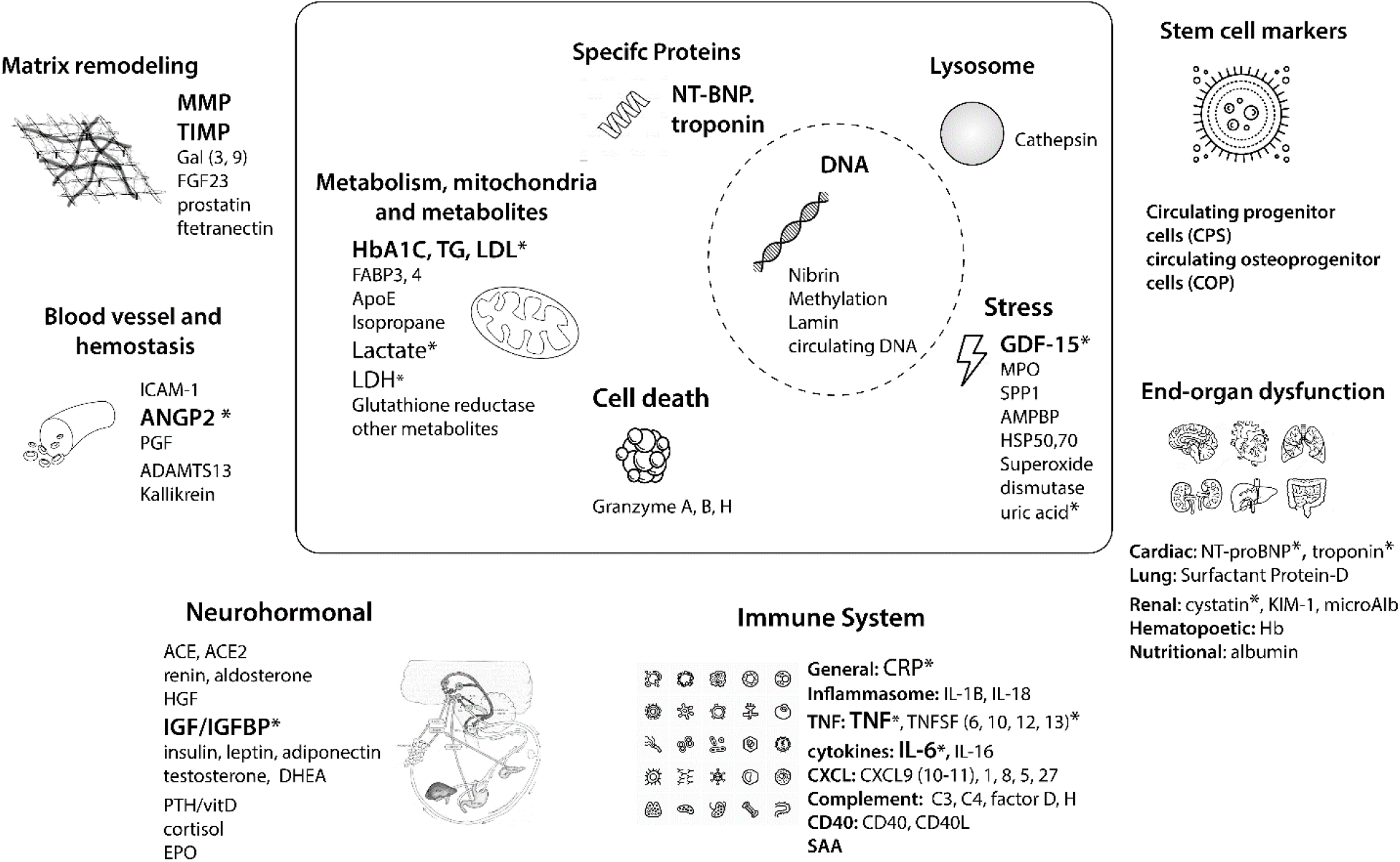
Schematic diagram of target biomarkers curated from literature.

**Figure 2.**
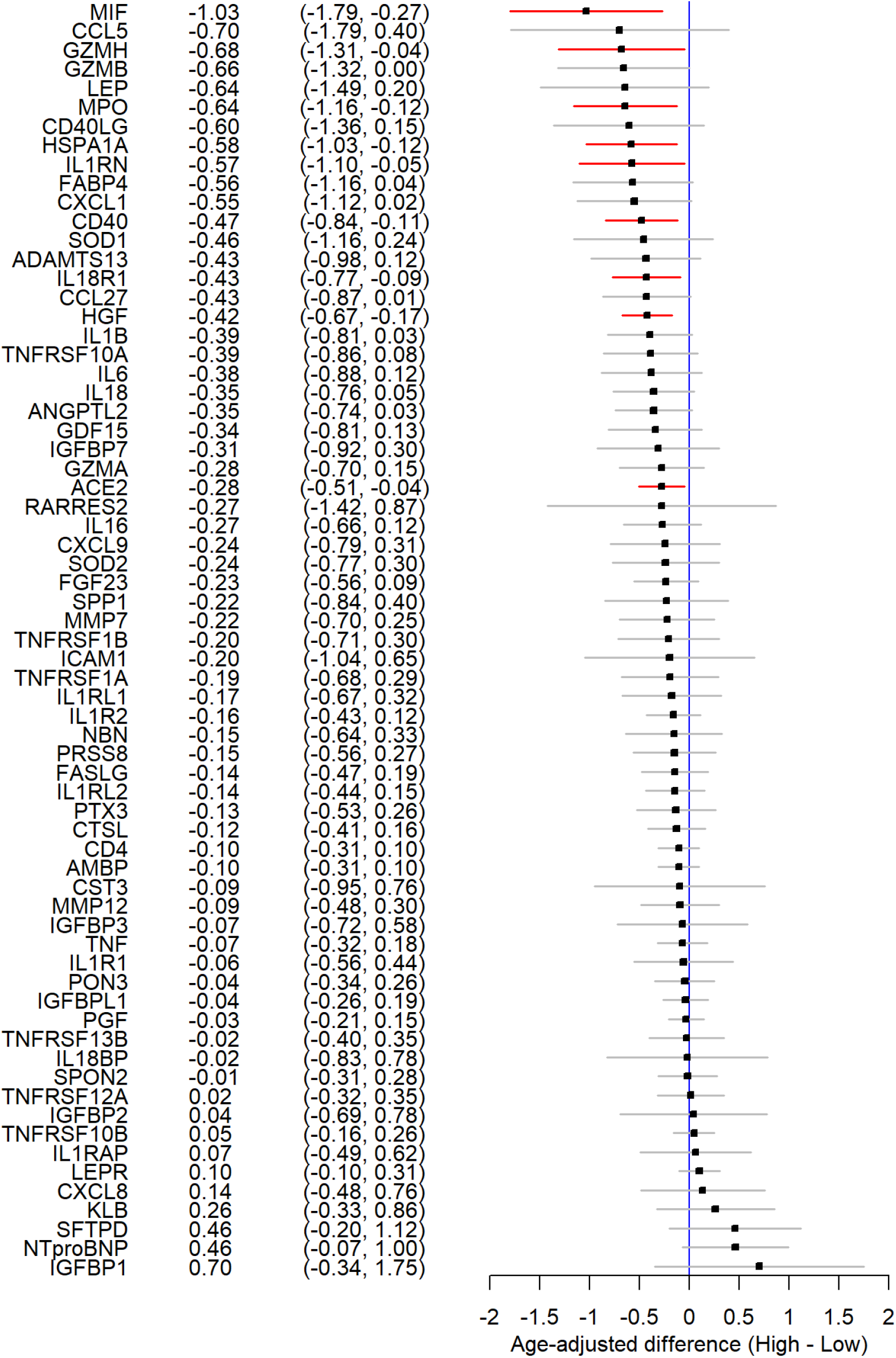
Age-adjusted differences with 95% confidence intervals in biomarker levels between high and low exposure groups. Markers with significantly lower relative abundance in the high exposure group are highlighted in red.

Between study visits (generally from lower to higher exposure to air pollution), firefighters witnessed significant increases in two biomarkers (MMP7 and NT-proBNP) and significant decreases in 11, including MIF and MPO. Unmatched controls witnessed significant increases in five biomarkers (nibrin [NBN]; granzymes A, B, and H; interleukin-16 [IL16]) and significant decreases in six, including MMP7 and C-C motif chemokine ligand 5 (CCL5).

For the inter-plate comparisons on the 10 samples processed on different plates, the median (interquartile range) of estimated Spearman’s rank correlation coefficients, overall and among target biomarkers, were 0.70 (0.45 – 0.89) and 0.66 (0.47 – 0.81), respectively. These summaries indicate that most associations were in the moderate to strong range.

## Discussion

Our pilot study following a major wildfire season demonstrated the feasibility of timely participant recruitment and plasma sample processing using targeted proteomics, as well as the application of novel statistical approaches to high-throughput data. Notably, the LASSO’s penalty-based selection algorithm allowed us to concentrate future validation efforts on the top 2% of nearly 1500 assays.

Three biomarkers from inflammation and oncology panels were found to be higher in firefighters compared to matched controls. Prior studies have noted the roles of ANGPT1 upregulation in wound-healing^16^ and of HAGH in the detoxification of reactive 2-oxoaldehydes.^17^ Upregulation of MMP1, which enhances the migration of tumor cells and adversely associated with tumor outcome, has been implicated in 11 cancer types. ^18^ Evidence also suggests that members of the MMP family are involved earlier in tumorigenesis, i.e., malignant transformation, angiogenesis, and tumor growth at both primary and metastatic sites.^19^

Among targeted biomarkers identified in secondary analyses to be numerically higher in firefighters with high exposure, NT-proBNP is a known prognostic indicator in chronic or acute heart failure^20^, while SFTPD regulates inflammation at pulmonary sites and is a causal risk factor of chronic obstructive pulmonary disease.^21^ Several biomarkers found to have increased with pollution exposure among firefighters and unmatched controls (MMP7,^22^ NBN,^23^ granzymes,^24^ and IL16^25^) have known roles in inflammation across multiple organ systems.

We anticipated that markers of the inflammasome pathway would be elevated in firefighters, especially among those with high exposure. To minimize confounding effects from the healthy worker effect (HWE), we carefully screened control participants to exclude comorbidities and ensure comparability.^26^ Reassuringly, leptin levels—known to be inversely associated with physical fitness—did not significantly differ between matched participants, although firefighters showed slightly lower levels on average compared to controls. Among firefighters, those with higher exposure had numerically lower leptin levels than their lower-exposure counterparts. This could be due to differences in fitness or adaptation to repeat exposure. These observations highlight the inherent challenges of cross-sectional studies involving firefighters (who generally have higher fitness levels and potentially better metabolic health than the general population) and suggest the most appropriate study design to be longitudinal. In addition, controlling for fasting state, time of day of sampling, and activity level in future studies would minimize confounding.

After a five-year data collection effort, the Occupational Requirements Survey from the Bureau of Labor Statistics recently identified the firefighting occupation as the most physically demanding among nearly 500 U.S. jobs.^27^ Despite this high level of physical fitness, comparisons between firefighters and matched controls, as well as between high- and low-exposure groups, revealed biomarkers associated with adverse health effects. These findings suggest that exceptional physical fitness alone may not provide sufficient protection against the health impacts of air pollution.

In future studies, longer follow-up with additional sample collection time points during and outside of wildfire season could establish within-participant biomarker changes and inform the degree of recovery or chronic pathology between exposures to smoke. Additionally, subgroup analyses conducted on a larger and more diverse cohort will provide insights into proteomic variations within the exposed population, especially among subgroups more vulnerable to PM_2.5._

## Abbreviations

ANGPT: angiopoietin
CCL5: C-C motif chemokine ligand 5
FDR: false discovery rate
HAGH: hydroxyacylglutathione hydrolase
HWE: healthy worker effect
IGFBP: insulin-like growth factor binding protein
IL16: interleukin-16
LASSO: least absolute shrinkage and selection operator
MIF: macrophage migration inhibitory factor
MMP: matrix metalloproteinase
MPO: myeloperoxidase
NBN: nibrin
NT-proBNP: N-terminal pro-brain natriuretic peptide
PLS-DA: partial least squares discriminant analysis
SD: standard deviation
SFTPD: surfactant protein D
VIP: variable importance in projection

## Acknowledgements

We would like to express our gratitude to Ms. Youn Soo Jung and Dr. John Coller for data curation and Dr. Yann Le Guen for methodological support. We also appreciate the intellectual contributions of Drs. Joseph Wu and PJ Utz toward the interpretation of findings.

## Author Contributions

K.N. and M.J. conceptualized and designed the study. All authors contributed to the acquisition, analysis, or interpretation of participant data. V.D. and Y.J. drafted the manuscript. All authors read, revised, and approved the final manuscript.

## Ethics Approval

This study was conducted according to the guidelines of the Declaration of Helsinki and approved by Stanford’s Institutional Review Board (Protocol IRB-8629).

## Data Availability

The datasets analyzed during the present study are available from the corresponding author upon reasonable request.

